# Uptake and Determinants of Hepatitis B Vaccination among Health Laboratory Practitioners in Tertiary Hospitals in Dar es Salaam, Tanzania: A Cross-Sectional Study

**DOI:** 10.64898/2025.12.03.25341607

**Authors:** Seleman Said, Mary Migiro, Dominic Renatus, Raidah R Gangji, Lilian Nkinda, Upendo Kibwana, Frank Msafiri, Doreen Kamori, Agricola Joachim, Joel Manyahi, Mtebe V Majigo, Salim S. Masoud

## Abstract

Hepatitis B virus (HBV) infection is a significant occupational risk for laboratory practitioners. Despite proven vaccine effectiveness and global recommendations, HBV vaccine uptake among healthcare workers in Tanzania remains low. This study assessed HBV vaccination uptake and its determinants among laboratory practitioners in tertiary hospitals in Dar es Salaam. An analytical cross-sectional study was conducted from March to June 2025 across four tertiary hospitals, enrolling 130 participants using the Kish-Leslie formula. Data were collected using a structured, pre-tested, interviewer-administered questionnaire and verified vaccination records, and analyzed with IBM SPSS v27. Descriptive statistics summarized participant characteristics, and associations were assessed using chi-square, Fisher’s exact tests, and Poisson regression with robust variance. Of 130 participants (median age 30 years, IQR: 25–36), 54.6% (71/130) completed the three-dose hepatitis B vaccination schedule, 23.8% (31/130) received partial doses, and 21.5% (28/130) were unvaccinated. Among the partially vaccinated, missed appointments (48.4%, 15/31) and lack of vaccine availability (41.9%, 13/31) were the main reasons. Half of the unvaccinated (50.0%, 14/28) cited lack of opportunity. Most participants knew the vaccine is essential (99.2%, 129/130), and 94.1% (96/102) acknowledged three doses are required for full protection. Participants with >2 years of work experience were nearly twice as likely to be vaccinated (aPR = 1.998, 95% CI: 1.064–3.173, p = 0.031). Those perceiving the vaccine as very or slightly effective were 44.8% (aPR = 0.552, 95% CI: 0.417–0.731, p = 0.010) and 41.9% (aPR = 0.384, 95% CI: 0.384–0.879, p < 0.001) less likely, respectively, to be vaccinated compared to those uncertain. HBV vaccine uptake among laboratory practitioners in Dar es Salaam is suboptimal, mainly due to structural barriers. Strengthening workplace vaccination programs, ensuring consistent vaccine supply, and implementing reminder systems could improve healthcare worker protection.

## Introduction

Hepatitis B virus (HBV) infection continues to be a significant concern for public health worldwide and a leading contributor to illness and death, especially in resource-limited countries [1,2]. The World Health Organization (WHO) estimated 257 million people were living with chronic HBV infection in 2022, which resulted in 1.1 million deaths worldwide [3,4]. HBV infection can lead to severe liver disorders such as liver cirrhosis and hepatocellular carcinoma [5,6].

Due to the increased rate of HBV infection and the worry for the world’s public health, the WHO developed a global viral hepatitis strategy in 2016 for the elimination of HBV infection by the year 2030. The strategy aimed at reducing new infections by 90% and the death rate by 65% [7]. In addressing viral hepatitis as part of the global health scope framework, an overarching health goal of the 2030 Agenda for Sustainable Development, Tanzania imposed a strategic plan 2018/19 – 2022/23 to manage and control viral hepatitis nationally, which recommends Hepatitis B vaccination in all healthcare workers (HCWs) [5]. According to WHO estimates, vaccination coverage among HCWs in countries with low- and moderate-income levels ranges from 18% to 39%, compared to 67% to 79% in high-income countries [8]. In Tanzania, the proportion of Hepatitis B vaccination coverage among HCWs ranges from 18.9 to 70.5% [9–11].

Among HCWs, laboratory practitioners represent a subgroup at especially high risk for occupational exposure to bloodborne pathogens, including HBV, due to frequent handling of blood specimens, sharps, and contaminated materials [12,13]. However, many studies on HBV vaccination and seroprotection tend to present aggregated data for HCWs generally, with limited disaggregation by laboratory role. Where such data are available, vaccination uptake among laboratory personnel has been shown to be lower than among physicians. For example, a cross-sectional study in Ethiopia reported HBV vaccination coverage of 41% among laboratory staff compared to 65% among physicians [14,15]. In Nigeria, vaccination uptake among medical laboratory scientists was similarly low, with only 32% fully vaccinated, despite high awareness of occupational risk [16,17].

Despite the introduction of the hepatitis B vaccination in Tanzania in 2002, vaccination coverage among HCWs remains suboptimal, with significant regional disparities and inadequate understanding of the barriers to vaccination [10,11]. This raises concerns about the potential risks of HBV transmission in healthcare settings and highlights the need for targeted interventions to improve vaccination rates. Therefore, in view of that, this study was designed to assess hepatitis B vaccination uptake and its predicting factors among healthcare laboratory practitioners working in the tertiary hospitals in Dar es Salaam.

## Materials and methods

### Ethics statement

This study was approved by the Institutional Review Board of Muhimbili University of Health and Allied Sciences (Ref.No.DA.282/298/01.C/2568). Permission to conduct the study was obtained from the respective hospital administrations. Before registration in the study, informed consent was obtained from all participants.

### Study design and setting

An analytical cross-sectional study was conducted between 21/3/2025 and 4/6/2025 across four tertiary hospitals in Dar es Salaam, Tanzania: Amana Regional Referral Hospital, Mwananyamala Regional Referral Hospital, Temeke Regional Referral Hospital, and Muhimbili National Hospital. Amana Regional Referral Hospital, serving the Ilala Municipality with approximately 1.2 million residents, operates with around 362 authorized beds and manages between 800 and 1,200 outpatient visits daily. Mwananyamala Regional Referral Hospital serves as the referral center for Kinondoni, Ubungo, and surrounding areas, covering a catchment population of over 2.2 million people. Temeke Regional Referral Hospital serves the Temeke District, which has a population of approximately 1.2 million and currently operates with 304 beds. Finally, Muhimbili National Hospital (MNH), the national referral, teaching, and research hospital, provides services to patients from all over Tanzania and neighboring countries, boasting a robust infrastructure with 1,500 beds. These hospitals were selected not only because they have a relatively higher number of laboratory workers compared to other facilities, but also because they provide a representative mix of regional referral and national hospital settings in Dar es Salaam.

### Study population

The study participants included laboratory practitioners working in clinical diagnostic laboratories at four selected hospitals. These practitioners consisted of laboratory technologists, scientists, and laboratory assistants who were actively involved in sample collection, processing, analysis, and reporting laboratory results. To qualify, participants had to provide written informed consent and verifiable hepatitis B vaccination records, such as a vaccination card. We excluded individuals who refused to participate or claimed to have received a hepatitis B vaccination without providing proof.

### Sample size and sampling technique

The minimum sample size of 130 participants was determined using Kish Leslie’s formula for a single population proportion, assuming a proportion p = 0.7 based on previous estimates of hepatitis B vaccination coverage among healthcare workers done at Kilimanjaro Christian Medical Center in 2021 [9], with a 95% confidence level, and an 8% margin of error. Probability proportional to size (PPS) was used to allocate the overall sample (n = 130) to the four hospitals in proportion to each hospital’s estimated laboratory practitioner population (Muhimbili 177, Amana 32, Temeke 28, Mwananyamala 30; total = 267). This site targets Muhimbili (86), Amana (15), Temeke (14), and Mwananyamala (15). Within each hospital, consecutive convenience sampling was applied, whereby participants were recruited as they came into contact with the research team until the required sample size was reached. Due to variations in staff availability and response rates across sites, the final recruited numbers differed from the PPS targets (Muhimbili: 76, Amana: 29, Temeke: 18, Mwananyamala: 7). These deviations from the PPS targets reflect practical field realities and did not impact the overall sample size required for analysis.

### Data collection

Data were collected using a validated structured questionnaire designed to capture relevant information from participants. The questionnaire was digitalized using Google Forms before the main data collection. The tool was pilot tested on 13 participants to assess clarity and functionality, and necessary adjustments were made to ensure data quality. Verbal consent was obtained before distributing the private Google Form link. The Google form link also contained a detailed written informed consent document before participants could participate in the study.

We also collected information on participants’ knowledge of hepatitis B vaccine effectiveness, number of recommended doses, safety, and occupational exposure risk, which was assessed using four separate questions in the structured questionnaire. For vaccine effectiveness, participants who responded, “very effective” were awarded 1 point, while responses of “less effective,” “slightly effective,” or “I don’t know” were scored 0. For vaccine safety, the correct response was “yes,” scored as 1 point, while “no” was scored 0. For occupational exposure risk, participants selecting “high risk” were awarded 1 point, while “low risk,” “moderate risk,” or “I don’t know” were scored 0. For knowledge on the number of recommended doses, the correct answer “three doses” awarded 1 point, while responses “two doses” or “one dose” were scored 0. These binary scores contributed to the overall knowledge score used in the analysis.

### Statistics

Data were analyzed using IBM SPSS Statistics version 27.0. Categorical variables were summarized as frequencies and percentages, while continuous variables such as age were presented as medians with interquartile ranges (IQR). The relationship between independent variables and hepatitis B vaccination uptake was assessed using the Chi-square and Fisher’s exact tests. To further determine the strength and direction of associations while controlling for potential confounders, Poisson regression with robust variance estimation was employed to calculate crude and adjusted prevalence ratios (cPR and aPR), along with corresponding 95% confidence intervals, in both univariate and multivariate analyses. A p-value of <0.05 was considered statistically significant.

Both statistical and epidemiological considerations guided the selection of variables for the multivariable analysis. Variables with a p-value less than 0.20 in univariate analysis were considered for inclusion, and those recognized in previous literature as potential confounders were also retained. In addition, age group and work experience were included a priori due to their theoretical relevance to both exposure and outcome, thereby ensuring adequate adjustment and reducing the risk of residual confounding.

## Results

### Demographic and professional characteristics

Of the 130 participants enrolled, 57.7% (75/130) were male, and the median age was 30 years, with an interquartile range (IQR) of 25–36 years. Most participants, 47.7% (62/130), were aged 40 years or older. The majority were from MNH, comprising 58.5% of participants (76/130). The microbiology and phlebotomy sections were the most represented, each with 19.2% (25/130).

Additionally, 81.5% (106/130) of participants had more than two years of work experience. Most respondents perceived themselves to be at high risk of HBV infection, 76.9% (100/130). Most participants believed the HBV vaccine to be very effective, 76.2% (99/130), and similarly, the vaccine was perceived as safe by 87.7% (114/130) of respondents. Nearly all participants, 99.2% (129/130), acknowledged the importance of the HBV vaccine for the healthcare workers “Table 1”.

**Table 1:** Proportion of hepatitis B vaccination uptake among health laboratory practitioners at Referrals and Tertiary Hospitals in Dar es Salaam.

| Variable | Total<br>*n* (%) * | Vaccination Uptake | P-value |
| --- | --- | --- | --- |
|  |  | Yes, *n* (%) |  |
| **Number of shots** |  |  |  |
| Complete | 71 (54.6) |  |  |
| Incomplete | 31 (23.8) |  |  |
| Not received any shot | 28 (21.5) |  |  |
| **Sex** |  |  | 0.077 |
| Male | 75 (57.7) | 36 (48.0) |  |
| Female | 55 (42.3) | 35 (63.6) |  |
| **Median age (IQR)** | 30 (25-36) | 30 IQR (26-36) | 0.099 |
| **Age group (years)** |  |  | 0.143 |
| 20-29 | 22 (16.9) | 16 (72.7) |  |
| 30-39 | 46 (35.4) | 25 (54.3) |  |
| 40+ | 62 (47.7) | 30 (48.4) |  |
| **Facility** |  |  | 0.777 |
| Muhimbili National Hospital | 76 (58.5) | 40 (52.6) |  |
| Amana Regional Referral Hospital | 29 (22.3) | 17 (58.6) |  |
| Temeke Regional Referral Hospital | 18 (13.8) | 9 (50.0) |  |
| Mwananyamala Regional Referral Hospital | 7 (5.4) | 5 (71.4) |  |
| **Laboratory section** |  |  | 0.454 |
| Phlebotomy | 25 (19.2) | 12 (48.0) |  |
| Hematology | 23 (17.7) | 15 (65.2) |  |
| Microbiology | 25 (19.2) | 16 (64.0) |  |
| B/ transfusion | 14 (10.8) | 7 (50.0) |  |
| Chemistry | 12 (9.2) | 5 (41.7) |  |
| Serology | 9 (6.9) | 6 (66.7) |  |
| Parasitology | 12 (9.2) | 6 (50.0) |  |
| Histology | 6 (4.6) | 1 (16.7) |  |
| Others | 4 (3.1) | 3 (75.0) |  |
| **Work experience (years)** |  |  | 0.006 |
| Less than 2 years | 24 (18.5) | 7 (29.2) |  |
| More than 2 years | 106 (81.5) | 64 (60.4) |  |
| **Perception on exposure risk toward HBV infection** |  |  | 0.067 |
| High risk | 100 (76.9) | 59 (59.0) |  |
| Low risk | 30 (23.1) | 12 (40.0) |  |
| **Perception on vaccine effectiveness** |  |  | 0.037 |
| Slightly effective | 24 (18.5) | 13 (54.2) |  |
| Very effective | 99 (76.2) | 51 (51.5) |  |
| I don't know | 7 (5.3) | 7 (100.0) |  |
| **Perception of vaccine safety** |  |  | 0.351 |
| Safe | 114 (87.7) | 64 (87.7) |  |
| Not safe | 16 (12.3) | 7 (43.8) |  |
| **Importance of vaccine to healthcare workers** |  |  | 0.454 |
| Yes | 129 (99.2) | 71 (55.0) |  |
| No | 1 (0.8) | 0 (0.0) |  |

\* Percentages represent the frequency of participants in each category out of the total participants. The percentages in the vaccination uptake represent the proportion of participants vaccinated within each category (row percentages).

### Uptake of hepatitis B vaccination among health laboratory practitioners

The overall uptake of the hepatitis B vaccine among study participants was 54.6% (71/130). Those with more than two years of service showed a higher vaccine uptake (60.4%, 64/106) compared to their counterparts with less than two years of work experience (29.2%, 7/24) (p = 0.006). Participants who reported not knowing the effectiveness of the vaccine showed 100% uptake (7/7), in contrast to those who believed the vaccine to be highly effective, 51.5% (51/99) (p = 0.037).

Although not statistically significant, we observed that female participants tended to have higher vaccine uptake, 63.6% (35/55), compared to males, 48.0% (36/75) (p = 0.077). Similarly, participants who perceived themselves as being at high risk of HBV exposure had a higher uptake rate of 59.0% (59/100) compared to those perceiving themselves at low risk (40.0%, 12/30) (p = 0.067). No significant difference was found in vaccine uptake based on participants’ age, health facility, laboratory section, or perceptions of vaccine safety and importance among healthcare workers. “Table 1”

### Barriers to Hepatitis B Vaccination and Knowledge of Recommended Doses

Despite a high level of knowledge and positive perceptions, some participants remained unvaccinated or incompletely vaccinated. Among those who did not complete the vaccine series, the main reasons were missed appointments (48.4%, 15/31) and unavailability of the vaccine at their workplace (41.9%, 13/31). For those unvaccinated, the most common reason was a lack of opportunity to receive the vaccine, at 50% (14/28). Most participants were aware that the recommended hepatitis B vaccination schedule consists of three doses, as indicated by 94.1% (96/102) “Table 2”.

**Table 2:** Barriers to HBV Vaccination and Knowledge of Recommended Doses.

| Variables | Frequency (*n*) | Percentage (%) |
| --- | --- | --- |
| **Reasons for not completing full vaccination (*n*=31)** |  |  |
| Missed appointment | 15 | 48.4 |
| Not available in my working place | 13 | 41.9 |
| It is expensive can't afford | 3 | 9.7 |
| **Reasons for not being vaccinated (*n* = 28)** |  |  |
| Vaccine is not available in my working place | 4 | 14.3 |
| It is expensive | 2 | 7.1 |
| I'm concerned about side effects | 8 | 28.6 |
| I have not been offered the chance | 14 | 50.0 |
| **Number of recommended doses of Hepatitis B vaccine (*n*=102)** |  |  |
| Three doses | 96 | 94.1 |
| I don't know | 6 | 5.9 |

### Factors influencing hepatitis B vaccination

In univariate analysis, participants with more than two years of work experience were twice as likely to be vaccinated compared to those with less than two years of work experience (cPR = 2.070, 95% CI: 1.089–3.935, p = 0.026). Perception of vaccine effectiveness as slightly effective or very effective had 45.8% (cPR = 0.542, 95% CI: 0.375–0.783, p = 0.001) and 48.5% (cPR = 0.515, 95% CI: 0.426–0.624, p =<0.001) reduced likelihood of uptake compared to those who perceived they don’t know the effectiveness of the vaccine. Participants aged 40 and above had a 33.5% reduced likelihood of being vaccinated compared to those aged 20-29 (cPR = 0.665, 95% CI: 0.463-0.956, p = 0.028).

After adjusting for potential confounders, specifically sex and perception of exposure risk towards HBV infection, two factors remained independently associated with vaccination uptake. Participants with more than two years of work experience had a 98.8% higher chance of vaccination than those with less than two years of work experience (aPR = 1.998, 95% CI: 1.064–3.173, p = 0.031. Conversely, those who perceived the vaccine was very effective or slightly effective had 44.8% (aPR = 0.552, 95% CI: 0.417–0.731, *p* = 0.010) and 41.9% (aPR = 0.384, 95% CI: 0.384–0.879, *p* < 0.001), respectively reduced vaccination rate compared to those who perceived not knowing the vaccine’s effectiveness. “Table 3”

**Table 3:**
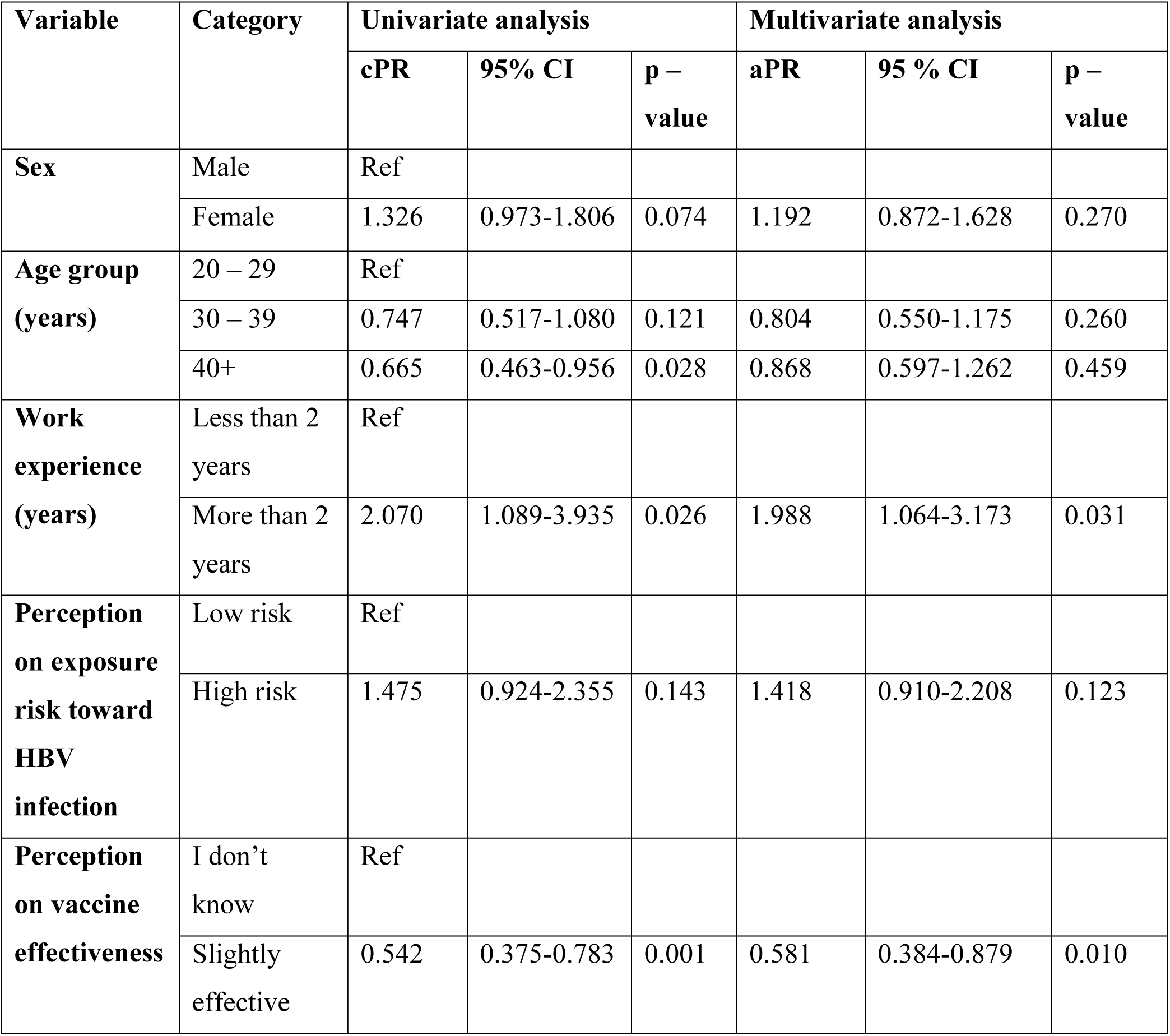
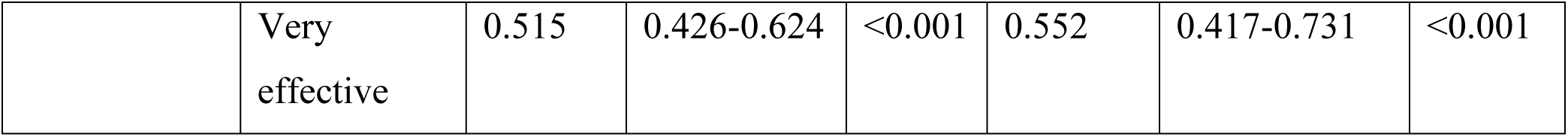
Univariate and multivariate analysis of factors influencing Hepatitis B vaccination uptake.

## Discussion

The results of this study reveal that while uptake of hepatitis B vaccination among health laboratory practitioners in Dar es Salaam shows progress compared to earlier Tanzanian and regional reports, uptake remains inadequate for full protection. The observed gaps in dose completion, coupled with structural barriers like vaccine unavailability and scheduling difficulties, highlight systemic weaknesses in occupational health programs. Interestingly, the counterintuitive association between strong belief in vaccine effectiveness and lower vaccination uptake suggests that access and programmatic issues, rather than knowledge deficits, may play a more decisive role in determining vaccination behavior.

In this study, the full-dose hepatitis B vaccine uptake among laboratory practitioners was 54.6%, which is relatively high compared to many reports from Tanzania and across Africa. A 2015 survey at Muhimbili National Hospital in Dar es Salaam reported 33.6% full vaccination and 56.9% with at least one dose, both lower than our estimate, suggesting gradual improvement over time [11]. More recent data from the same hospital in 2025 found 46.3% uptake, again slightly below our figure [18]. In contrast, a study from the Mwanza region reported only 18% of individuals were fully vaccinated, highlighting significant disparities between urban referral centers and peripheral facilities [6]. The high uptake in this study was due to the study’s focus on health laboratory practitioners in urban referral hospitals, who are routinely trained in biosafety and infection control. In contrast, other studies included broader HCWs, often with lower risk perception and weaker access to vaccination. Additionally, vaccination uptake may have increased over time due to national policies, heightened awareness of HBV, and occupational safety campaigns.

Regionally, the uptake in East Africa has historically been lower. For example, only 12.8% of Kenyan HCWs in a 2006 study had ever received the vaccine [19], and pooled estimates from Ethiopia place full-dose coverage at approximately 20%, with a recent survey of Addis Ababa nurses reporting 36.9% coverage [20,21]. In Uganda, mass vaccination campaigns achieved a 20% completion rate of the three-dose schedule, underscoring persistent challenges in community uptake [22]. Rwanda stands out as an exception: in a tertiary hospital cohort, nearly all participants who had initiated vaccination had completed all three doses, reflecting the impact of well-structured institutional programs [23].

At a broader continental level, a meta-analysis across Africa reported 24.7% full-dose coverage among HCWs, with the East African subgroup even lower at 15.1% [24]. Against this backdrop, our estimate of 54.6% uptake compares favorably, positioning our study population above both the national and regional averages. Nonetheless, coverage remains far from optimal, particularly in achieving universal protection, highlighting the need for more comprehensive Hepatitis B vaccination programs among healthcare workers in Tanzania.

Multivariable analysis revealed that two factors independently predicted hepatitis B vaccine uptake: work experience and perception of vaccine effectiveness. Healthcare workers with more than two years of service were significantly more likely to have been vaccinated compared with those with less than two years of experience, a finding consistent with other Tanzanian and regional studies that link longer occupational exposure to greater vaccination rates [6,11].

Interestingly, however, respondents who reported the vaccine as “very effective” or “slightly effective” were less likely to be vaccinated than those who responded, “don’t know.” This negative association contrasts with findings from Ethiopia and other African settings, where positive perceptions of vaccine effectiveness were generally associated with higher [24,23]. Globally, reviews also report that confidence in vaccine efficacy usually promotes uptake [26]. The divergence observed in our study may reflect contextual factors such as limited vaccine availability, programmatic barriers, or response bias, underscoring the need for qualitative inquiry to clarify how attitudes toward effectiveness translate into vaccination behavior in our setting.

The leading barriers to completing the hepatitis B vaccine series in our study were missed appointments (48.5%) and unavailability of vaccines at the workplace (41.9%). These findings are broadly consistent with a meta-analysis of African healthcare workers, in which 50.5% cited vaccine unavailability and 37.5% reported demanding work schedules as reasons for non-completion. In comparison, nearly half 50% had never been offered vaccination opportunities at all [24]. Tanzanian studies similarly highlight structural challenges, such as a lack of institutional vaccination programs and inconsistent vaccine supply, as major impediments [6,11]. Globally, systematic reviews concur with these findings, identifying poor access, limited workplace vaccination services, and weak reminder systems as recurring barriers to series completion among healthcare workers [26]. Collectively, this evidence highlights the importance of structured, facility-based immunization programs that ensure vaccine availability, integrate appointment reminders, and strengthen supply chain systems to enhance coverage and ensure completion of the full vaccine schedule.

This study provides valuable insight into hepatitis B vaccination coverage among laboratory practitioners in Dar es Salaam, contributing to the limited local data on occupational vaccine uptake. A key strength is the use of a structured data collection approach and verification of vaccination status, which enhances the accuracy of reported uptake. Additionally, including multiple hospitals improves the representativeness of the findings within the city’s healthcare setting. However, the use of convenience consecutive sampling within hospitals may introduce selection bias, potentially limiting the generalizability of the results.

## Conclusion

Although more than half of the participants achieved full-dose coverage, nearly half were either partially vaccinated or unvaccinated, leaving a significant proportion at risk of HBV infection. Work experience appears to promote uptake; however, programmatic obstacles, such as missed appointments and inconsistent vaccine supply, remain substantial barriers to uptake. To improve protection of healthcare workers, hospitals should prioritize on-site vaccination services, ensure reliable vaccine availability, and adopt reminder systems or mandatory pre-employment vaccination policies. These measures would close existing gaps and strengthen progress toward the elimination of HBV targets.

## Acknowledgements

The authors would like to express their sincere gratitude to the healthcare laboratory practitioners from Muhimbili National Hospital, Amana Regional Referral Hospital, Temeke Regional Referral Hospital, and Mwananyamala Regional Referral Hospital for their participation and cooperation in this study. We also extend our appreciation to the hospital management teams for granting permission and facilitating data collection. Special thanks go to the Department of Microbiology and Immunology at Muhimbili University of Health and Allied Sciences for providing technical guidance and academic support throughout the study.

## Author Contributions

SS, SM, JM, MVM, UK conceptualized the study. DR, RG, SS, MM were involved in acquisition of data. DK, MM, SS, MVM, performed statistical analysis and data interpretation. SS, SM, MM, DR drafted the manuscript. FM, LN, AJ, JM reviewed the original draft critically. All authors have read and agreed to the published version of the manuscript.

## Data Availability Statement

The datasets generated and/or analyzed during the current study are not publicly available due to confidentiality agreements with participants but are available from the corresponding author on reasonable request.

